## Supporting_Information_for_online_publication for "Strong isolation by distance and evidence of population microstructure reflect ongoing *Plasmodium falciparum* transmission in Zanzibar"

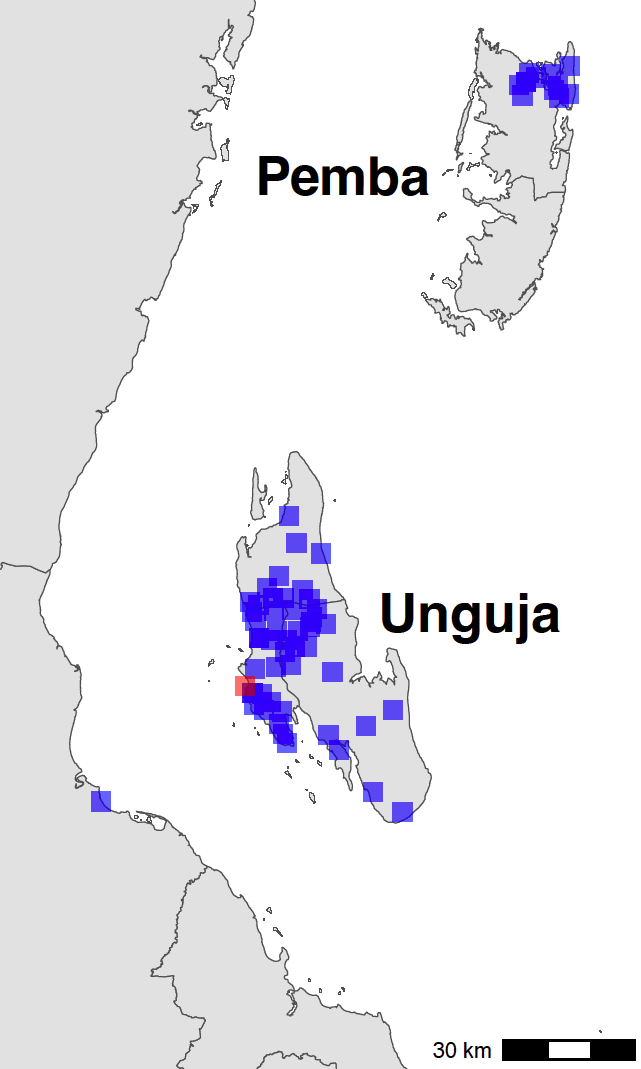

**Supplementary Figure 1. Sampling locations in Zanzibar (*shehia*) and Mainland (Bagamoyo district) Tanzania.** The centroids of the sampling locations are shown as blue rectangles. The ferry terminal in Zanzibar town in shown as a red rectangle. In Zanzibar, samples were collected throughout Unguja and in northern Pemba. In mainland Tanzania, samples were collected from Bagamoyo district.

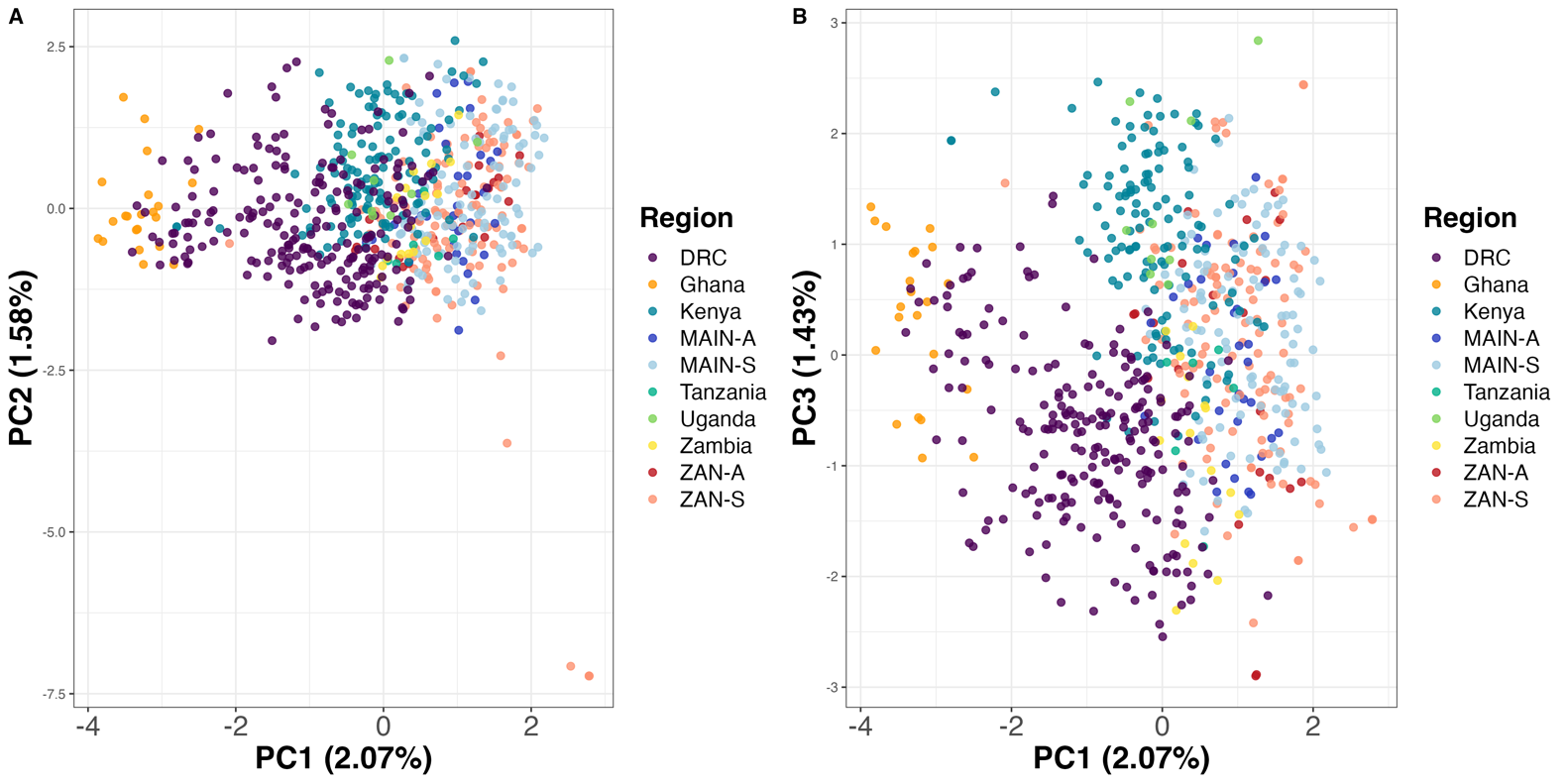
**Supplemental Figure 2. PCA utilizing samples across Africa show clustering based on geographic location.** Samples from Ahero, Kenya (n=147), a random 20% of samples from 5 regions across Africa (Verity et al. 2020) (n = 275) and from this study (n=282) were subsetted to 756 common loci. Within sample allele frequency (WSAF) was calculated, with an imputation step to replace missing values with the median WSAF, to perform PCA.

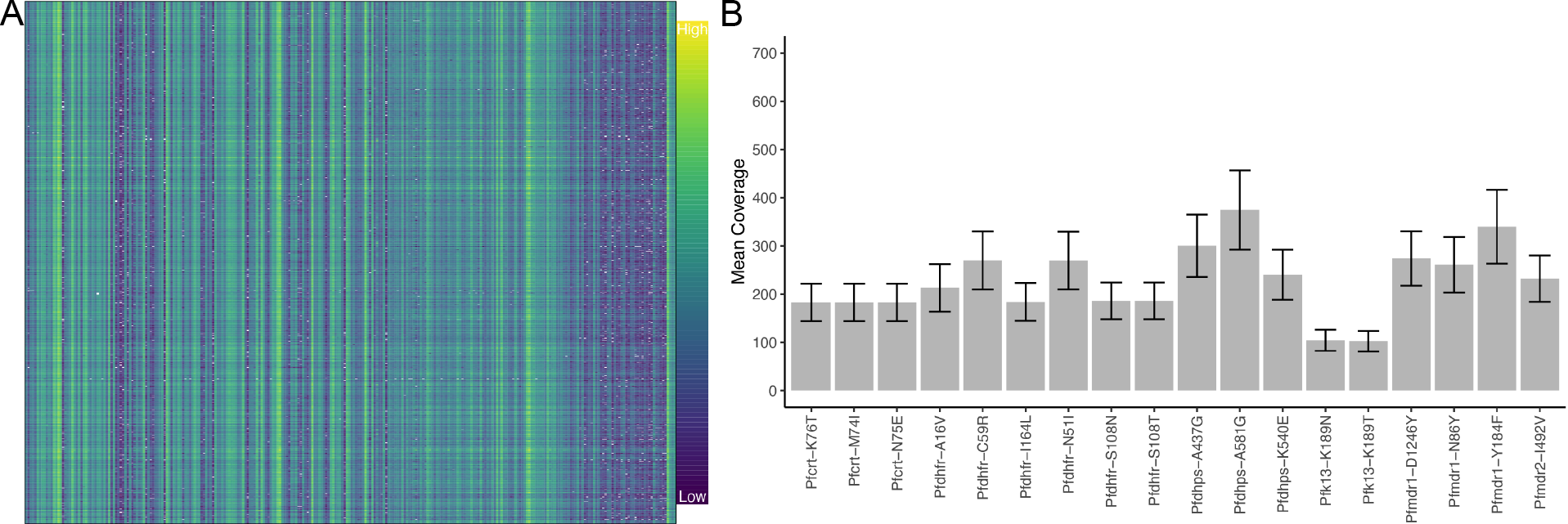

**Supplemental Figure 3. MIP performance shows coverage of loci.** Panel **A** shows the log transformed read depth for genome-wide SNPs for samples (columns) and loci (rows). The log transformed unique molecular identifier (UMI) count ranges from 0 to 9.87. Panel **B** shows the mean UMI coverage for the analyzed drug resistance mutations with a nonparametric bootstrap 95% CI.

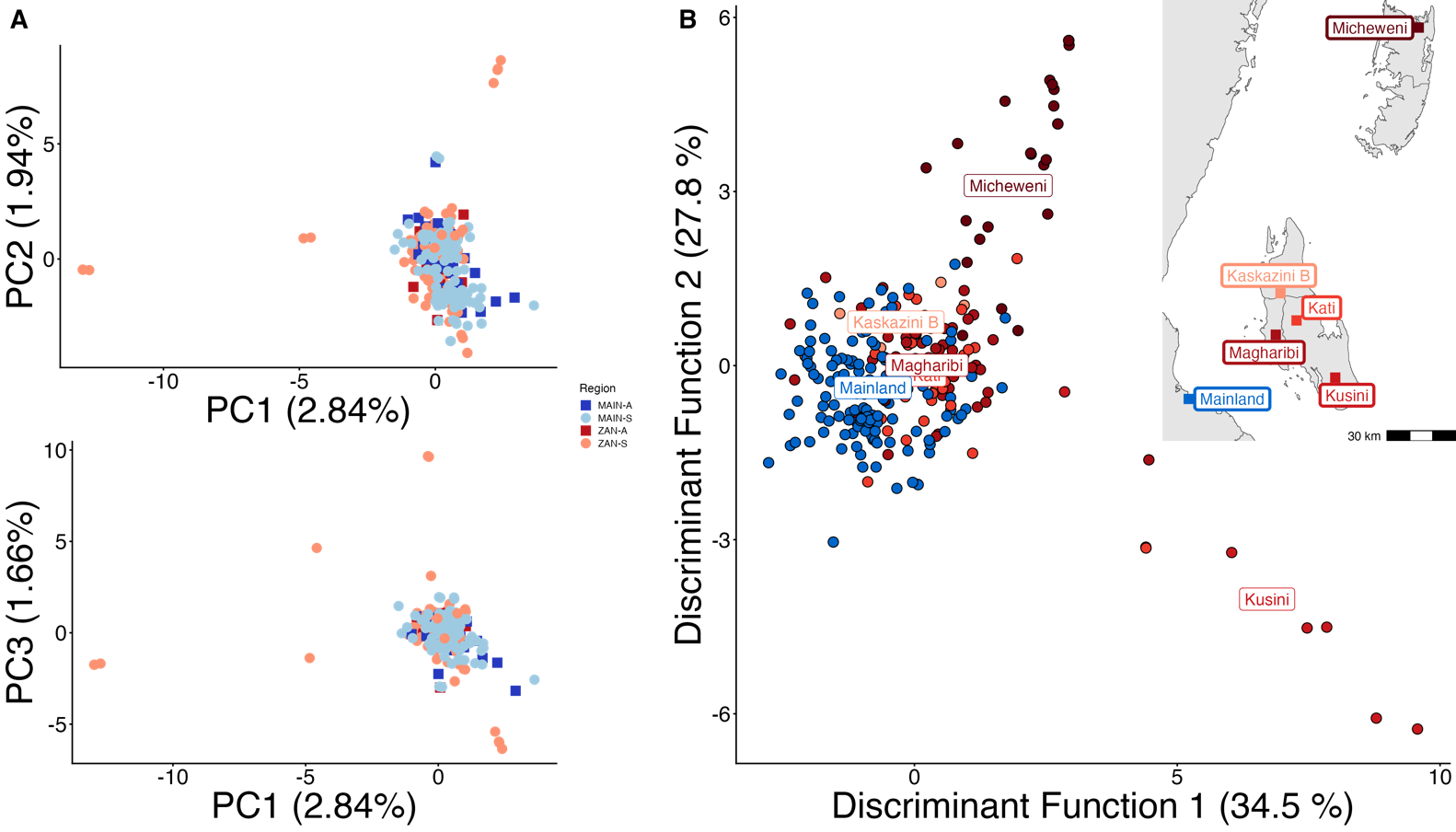

**Supplemental Figure 4. PCA with highly related samples shows population stratification radiating from coastal Mainland to Zanzibar.** PCA of 282 total samples was performed using whole sample allele frequency (**A**) and DAPC was performed after retaining samples with unique pseudohaplotypes in districts that had 5 or more samples present (**B**). As opposed to Figure 1, all isolates were used in this analysis and isolates with unique pseudohaplotypes were not pruned to a single representative infection.

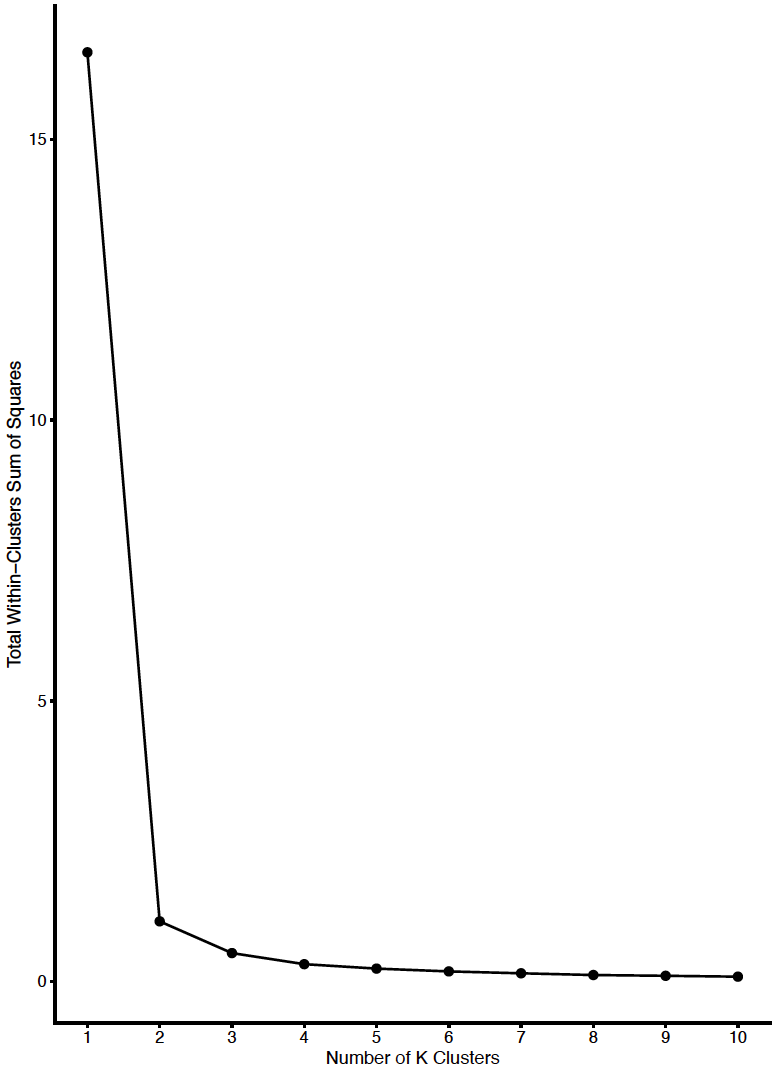

**Supplemental Figure 5. Diagnostic plot showing total within-cluster sum of squares versus number of clusters for determination of optimal K.** Mainland samples were considered an independent cluster. We selected a K of 4 for determining clusters on Zanzibar based on the inflection point above.

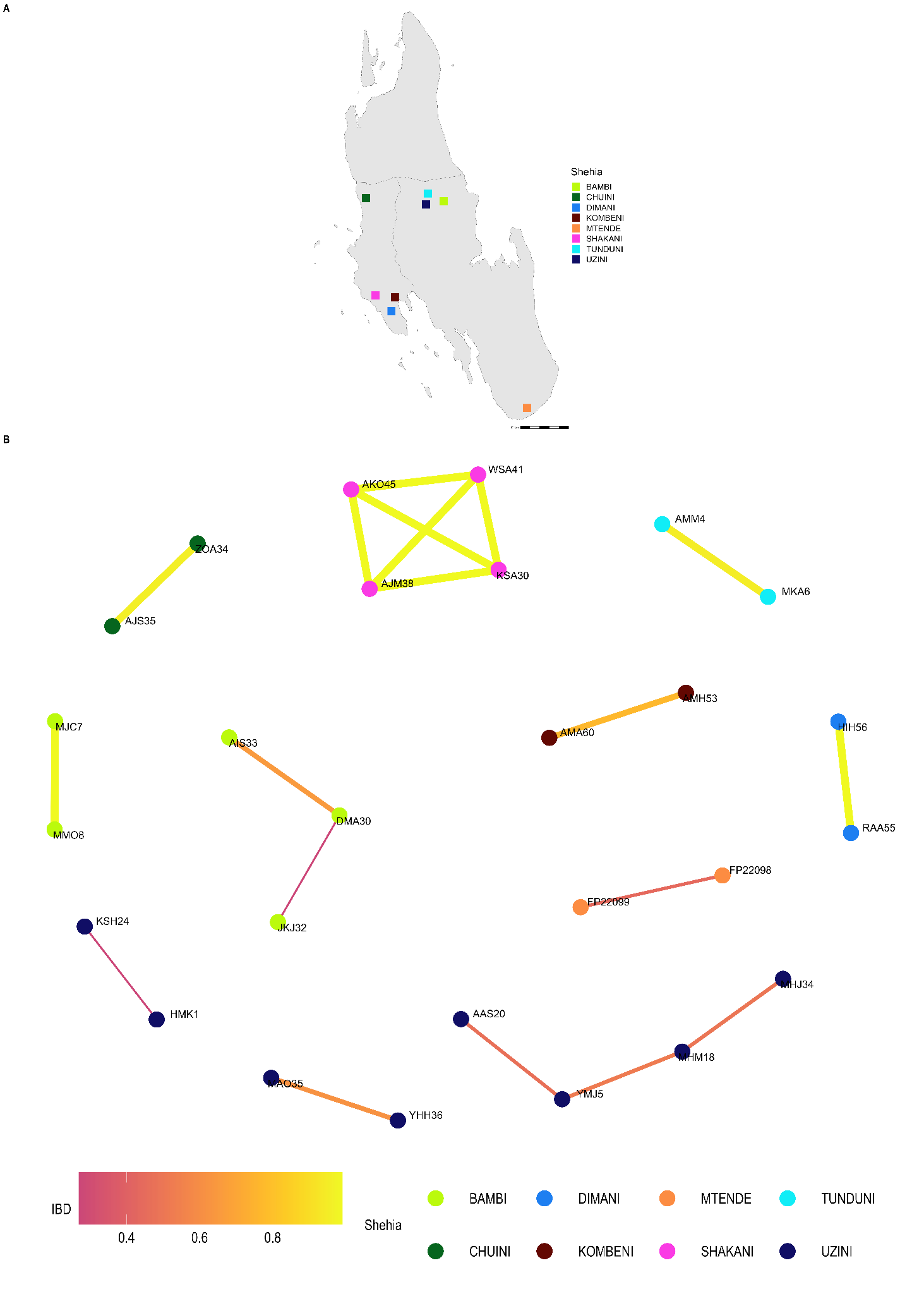

**Supplemental Figure 6: Network analysis of within *shehia* comparisons with an IBD of 0.25 or greater in Unguja.** Pairwise IBD comparisons of 0.25 or greater within different *shehias* were used. If a *shehia* is not represented, it does not have a pairwise comparison meeting the IBD threshold. *Shehias* that did contain a pair with an IBD of 0.25 or greater are plotted (**Panel A**). Network analysis of related pairs (IBD > 0.25) is plotted in **Panel B**. The width of each line represents higher magnitudes of IBD between pairs.

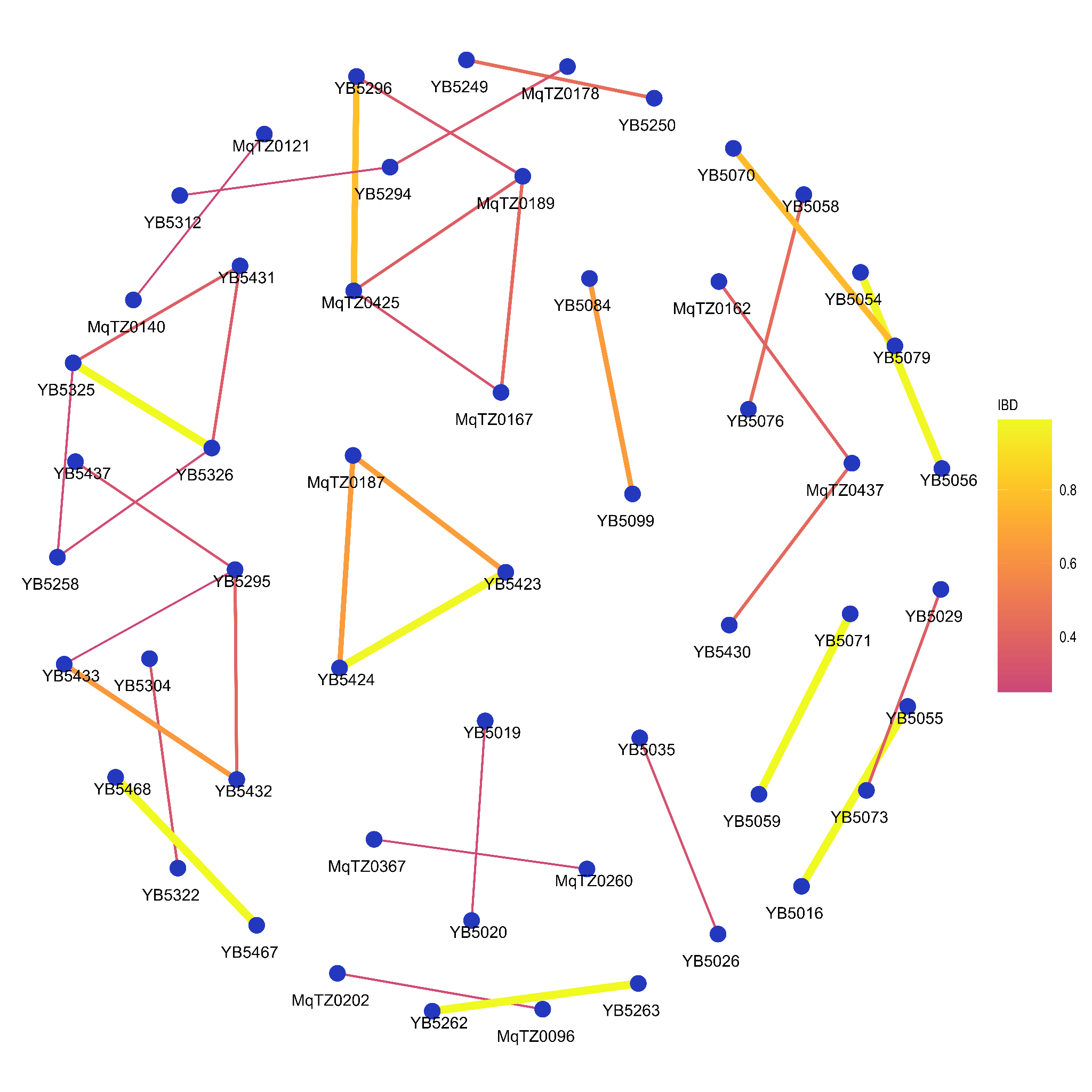

**Supplemental Figure 7: Network analysis of sample pairs with IBD of 0.25 or greater for coastal mainland Tanzania.** The network of highly related (IBD > 0.25) pairs is plotted above within coastal mainland Tanzania. The width of each line represents higher magnitudes of IBD between pairs.

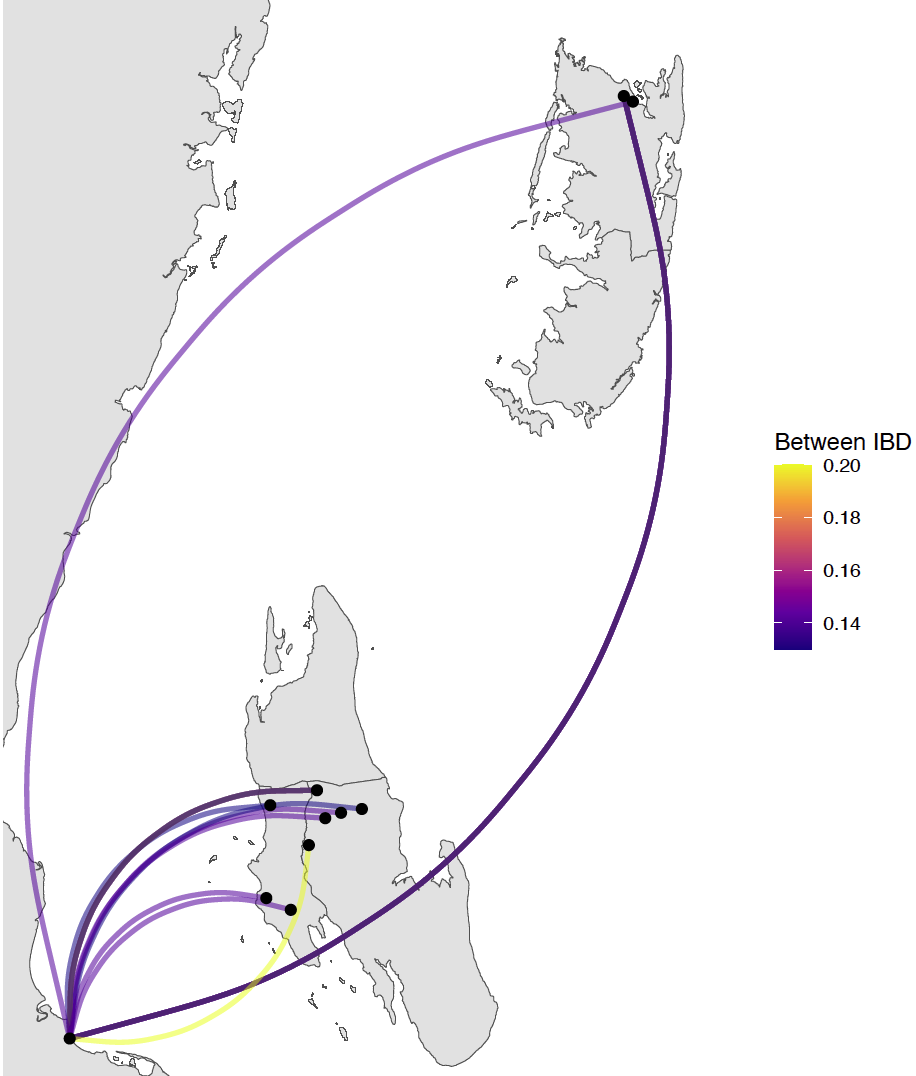

**Supplemental Figure 8. Sample pairs with an IBD of 0.125 or greater between Zanzibar and mainland Tanzania.**  Relatively few sample pairs showed moderate levels of IBD (between 0.125 and 0.20) between the coastal mainland and Zanzibar.

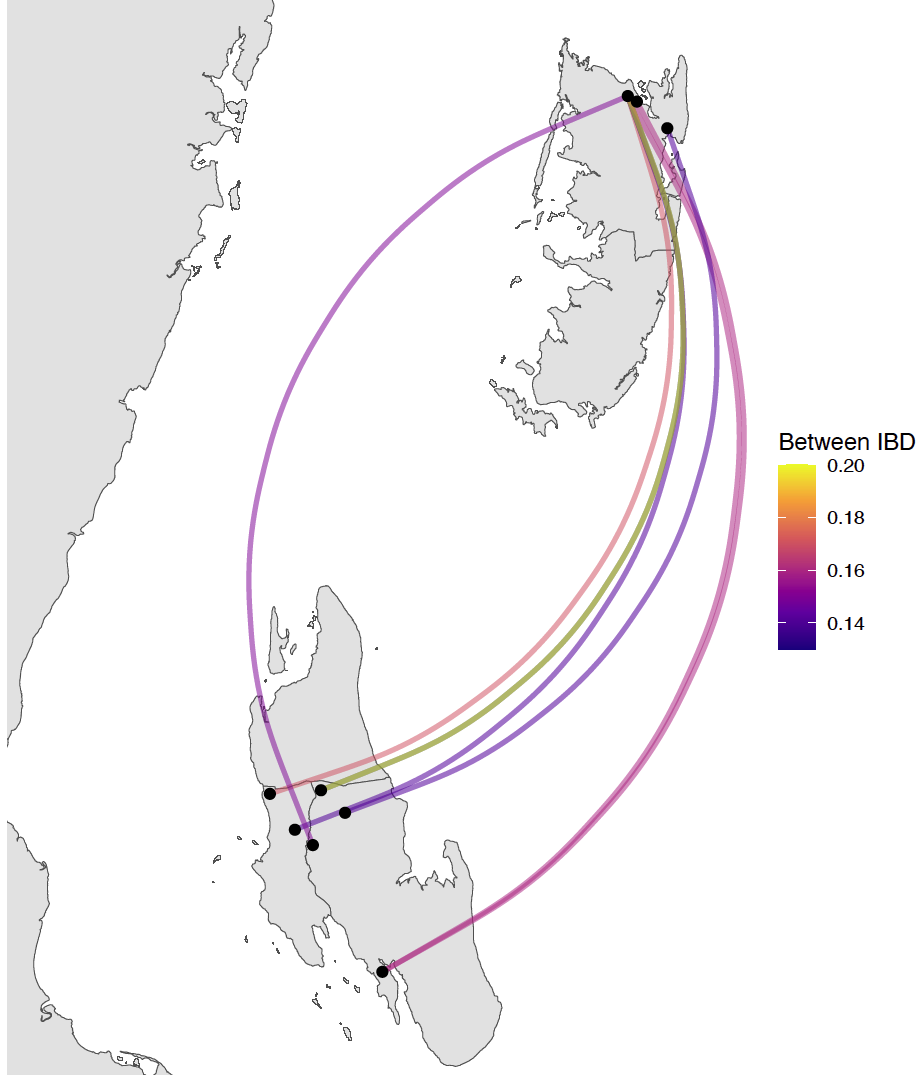

**Supplemental Figure 9. Sample pairs with IBD of 0.125 or greater between Unguja and Pemba.** Relatively few sample pairs showed moderate levels of IBD (between 0.125 and 0.20) between Unguja and Pemba.

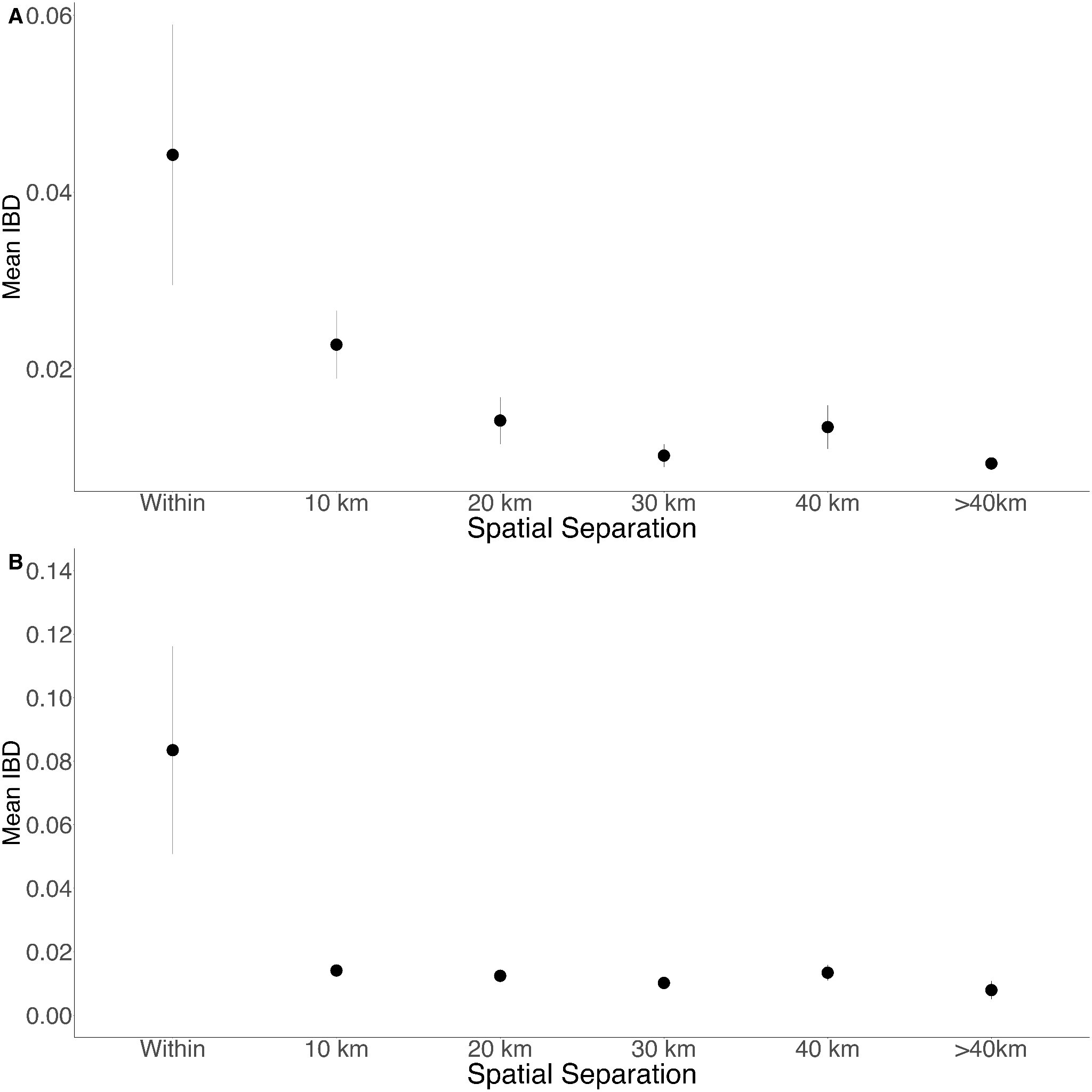

**Supplemental Figure 10. Isolation by distance in Zanzibar isolates (A) and only Unguja isolates (B).** Samples were filtered based on location and greater circle distance were calculated. These distances were binned at 10km increments.
